# Adapting a Shared Digital Communication Intervention for Caregivers of Children with Physical Disabilities and Professionals through Co-design: A Workshop Study

**DOI:** 10.64898/2026.11.10.26367483

**Authors:** Lili Worre Høpfner Jensen, Simon Kristoffer Johansen, Kristian Damgaard Lyng, Ole Rahbek, Søren Kold

## Abstract

**Introduction:** Caregivers of children with disabilities act as intermediaries between professionals due to fragmented communication and information sharing across sectors. The use of shared digital communication may support integrated care. Therefore, this study explored challenges and needs among caregivers and professionals and how an existing shared digital communication intervention could be adapted to support communication and coordination in this context.

**Methods:** Three future workshops involved 26 co-designers: caregivers (n=8), professionals across sectors (n=14), and software developers (n=4). Caregiver and professional data were analysed thematically, followed by matrix analysis to identify convergences, tensions, and design principles. In workshop with software developers, design principles were translated into design specifications.

**Results:** Both caregivers and professionals described how caregivers become de facto care coordinators. Clear responsibilities, continuity, and shared knowledge were identified as essential for improving communication and coordination. Shared digital communication was considered promising, but tensions emerged between accessibility and workload, and between transparency and confidentiality. The adapted intervention incorporated structured communication, shared responsibility, relational continuity, a shared overview, and a human digital coordination function.

**Discussion/conclusion:** The adapted intervention provides a foundation for cross-sector communication between caregivers and professionals and adds to the evidence base of communication solutions that may strengthen integrated care.

## 1. Introduction

Caring for a child with disability requires continuous involvement of professionals across health, social, and educational sectors, making effective communication and coordination essential. However, discontinuities in communication and limited information sharing often leave caregivers acting as intermediaries (Kouther et al., 2022; Pousada et al., 2013; Raina et al., 2005; Vadivelan et al., 2020). Limited caregiver access to professionals’ electronic records further exacerbates this challenge. Studies describe caregivers as “semi-professional coordinators”, adding to their substantial burden (Gulmans et al., 2009; Jensen et al., 2024a).

Communication and coordination also challenge professionals collaborating across disciplines and sectors (Jensen et al., 2024b; Styczen et al., 2025), and a key contributing factor is the fragmented organisation of services (Ghannam et al., 2025). Despite co-financing and close collaboration between regional and municipal levels, professionals operate under different legislative frameworks and use separate electronic records. This system fragmentation undermines integrated care in complex pathways requiring sustained cross-sector collaboration, and may increase caregiver burden by shifting responsibility for coordination onto families. Supportive and preventive integrated care interventions are therefore needed to strengthen collaboration across sectors and alleviate this burden.

In previous studies, we have explored a shared digital communication intervention connecting patients with healthcare professionals across sectors in an orthopaedic context. The benefits included real-time information sharing after surgery, interprofessional learning (Jensen et al., 2024a, 2024b), improved access to relevant professionals, and greater continuity of care around discharge (Jensen et al., 2024a). A randomised controlled trial also demonstrated that providing patients with access to the intervention significantly reduced hospital phone calls after discharge (Høpfner Jensen et al., 2024). However, complex interventions cannot automatically be assumed to be feasible or effective across context without adaptation (Moore et al., 2021a; Skivington et al., 2024, 2021). In the context of children with physical disabilities, the intervention would need to accommodate caregivers and a broader multidisciplinary network across health, social and educational sectors, including public and private providers, requiring changes to professional practices, workflows, and organisations. This complexity necessitates careful adaptation (Skivington et al., 2024, 2021).

Stirman et al. (2013) distinguish between adaptations to intervention content, including components, procedures, materials, and delivery methods, and adaptations to context, including implementation conditions, settings, formats, and delivery personnel (Stirman et al., 2013). Co-design can support adaptation of a complex intervention by identifying end users’ needs and practical challenges, such as workflow, communication, and organisational contraints (Vargas et al., 2022). It can also foster stakeholder ownership and commitment, supporting integration into routine practice and strengthening implementation and scalability (Moore et al., 2021a).

The aim of this workshop study was twofold. Firstly, we aimed to explore the challenges and needs related to communication and coordination among caregivers of children with physical disabilities and the professionals involved in the child’s care across sectors, and secondly, we aimed to adapt a shared digital communication intervention for this context through co-design.

## 2. Methods

### 2.1 Study Design

This study is a part of a larger research project with the overall aim of reducing the communication and coordination burden for caregivers of children with physical disabilities (SHARE IT project: shareitproject.dk). The project builds upon the framework for developing and evaluating complex interventions as defined by the British Medical Research Council (MRC), and involves four overall phases; adaptation of the intervention, feasibility testing, evaluation, and preparation for implementation (Skivington et al., 2024, 2021) (Figure 1).

**Figure 1.**
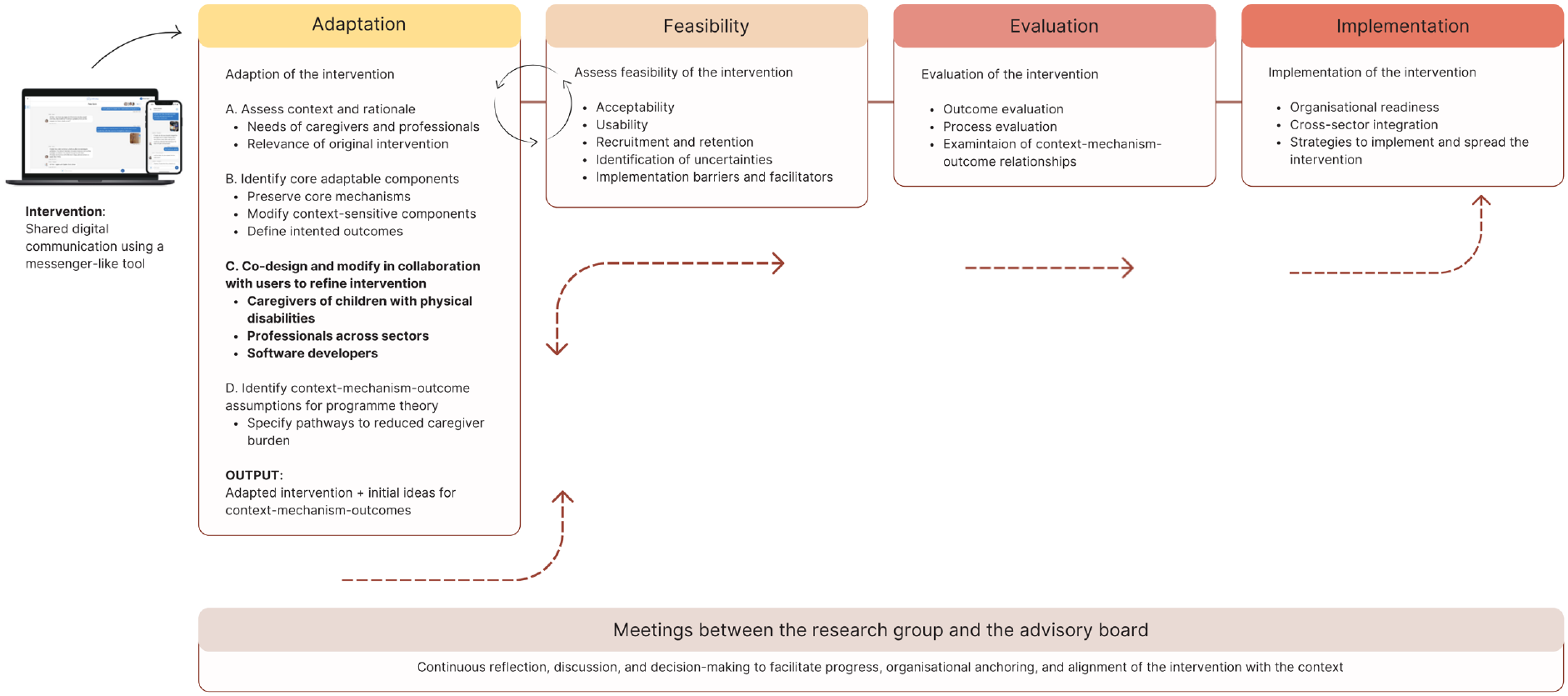
The four overall phases of the SHARE IT project include adaptation, feasibility, evaluation and implementation. The study reported in this paper is a part of the adaptation phase (highlighted with bold). During all phases of the project, the research group engages in discussions with the advisory board consisting of caregivers, researchers, clinicians, managers and legal experts. The advisory board also functions as an adaptation team and supports continuous reflection, and decision-making to facilitate progress, organisational anchoring, and alignment of the intervention with the contexts.

This study was informed by the ADAPT guidance for transferring complex interventions to new contexts (Moore et al., 2021a), and co-design as an early-phase collaborative process in which stakeholders jointly shape intervention development or adaptation (Vargas et al., 2022). We report on participatory workshops conducted with caregivers, professionals across health, social, and educational systems, and software developers to explore challenges and needs, and to co-design and adapt the intervention to support shared digital communication between caregivers and professionals.

The adaptation process was informed by the GUIDED framework to support a systematic and transparent approach to intervention adaptation, including stakeholder involvement, decision-making, and consideration of contextual factors (Duncan et al., 2020) (Appendix 1a). Adaptations to the intervention were subsequently documented and reported using the FRAME framework (Wiltsey Stirman et al., 2019) (Appendix 1b).

### 2.2 Setting

The workshops with caregivers and professionals were held at a conference venue close to Aalborg University Hospital on October 28, 2024 and January 29, 2025, respectively. This neutral setting was chosen instead of the hospital to avoid activating underlying power dynamics that could be counterproductive for the workshops (Trettin et al., 2025). The workshop with software developers took place at their facilities and was conducted March 3, 2025.

### 2.3 Participants as co-designers

In this study, we regard all participants as co-designers, recognizing their different knowledge and expertise as essential resources in the adaptation process (Vargas et al., 2022). Participants were sampled by convenience, based on availability, which is a common strategy in workshop-based research, particularly when engaging lead users, that are typically proactive and motivated to contribute, making them valuable for driving the development of novel product concepts (Trettin et al., 2025; von Hippel, 1986).

Caregivers were invited to participate by distributing flyers in the Department of Pediatrics and the Department of Orthopaedics at Aalborg University Hospital, and via social media. Participation was not limited to specific diagnoses or physical disabilities in the child as there may be comparable communication and coordination needs across groups.

Key inclusion criteria for caregivers were that the child was < 18 years and required assistance from and interacted with multiple institutions and professionals across sectors, such as hospitals, municipal departments, and/or specialized day care and school facilities. This ensured that the caregivers had experiences with the complex challenges that exist in cross-disciplinary and cross-sector communication and coordination.

Professionals were invited through phone calls to managers at various institutions in the North Denmark Region and sampled to represent different disciplines and sectors. Eligible professionals worked with children with physical disabilities in their daily practice and had more than two years of experience. This ensured, that they had sufficient knowledge of communication and coordination practices in the context of children with disability.

In the third workshop, employees from the company providing the communication tool for the intervention, participated. They represented different competencies including management, software developer, multimedia designer and account manager. This group is collectively referred to as software developers throughout the paper.

### 2.4 Data collection

The initial planning of the workshops was guided by discussions in the project group and advisory board, and data collection was performed in three consecutive workshops. While the first two workshops with caregivers and professionals focused on gaining deeper insights into their challenges and needs, validating the relevance of the intervention in the given context, and collaboratively developing design principles for adapting the intervention with end users, the third workshop focused on translating these design principles into concrete design specifications for the communication tool. Therefore, a period of time elapsed between the first two workshops and the third, during which we analysed the data. We anticipated that this process would contribute to the overall co-design of the intervention (Figure 2).

**Figure 2.**
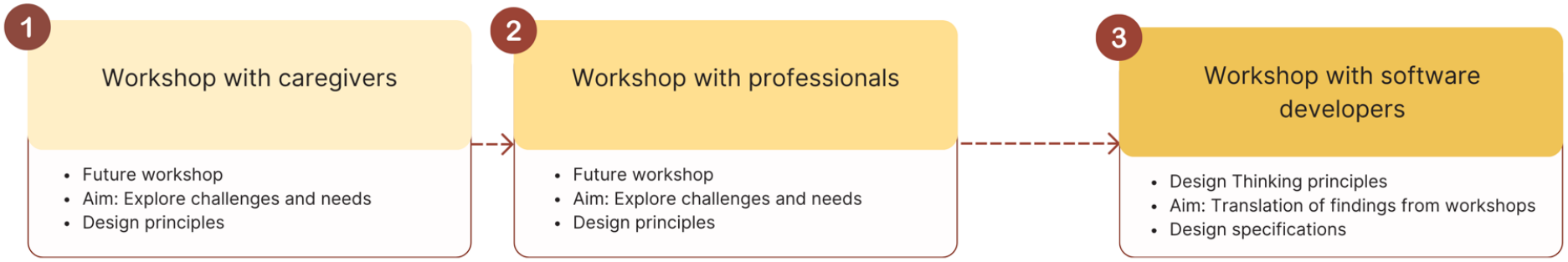
Data was collected through three workshops with co-designers: caregivers, professionals and software developers.

Details of the workshops were planned by LWHJ in collaboration with KDL and SKJ, who have previous experiences in planning and conducting workshops as a method for gathering qualitative user-experience data. LWHJ facilitated the workshops with assistance from KDL and SKJ. All workshops were audio recorded.

Structure of the workshops with caregivers and professionals

The workshop with caregivers and professionals followed the same structure in order to get comparable insights into their perceptions of challenges, needs and visions for the adaptation of the shared digital communication intervention. Both workshops were organized based on the principles of The Future Workshop (Jungk and Müllert, 1984) in three phases: a critique phase, a fantasy phase, and an implementation phase (Jungk and Müllert, 1984; Vidal, 2005) (Appendix 2) and using artifacts including a case, activity cards and posters to increase reflection and ideation (Appendix 3).

Caregivers and professionals were divided into groups of three to four individuals to make room for in-depth discussions. Groups were based on maximum diversity to ensure a broad range of perspectives, experiences, and competencies were represented within each group. In the workshop with caregivers this meant that caregivers were grouped with other caregivers who had children with different physical disabilities, and in the workshop with professionals, they were grouped with professionals from different disciplines and sectors. The workshops lasted approximately three hours each.

Structure of the workshop with software developers

The workshop was conducted using Design Thinking principles and guiding questions created by the concept of “How Might We” (Cross, 2011). Design Thinking is a human-centered, iterative problem-solving approach that focuses on understanding user needs to create innovative solutions that improve services, experiences, and outcomes. It was considered relevant for the purpose of the third workshop, as it emphasizes empathy, collaboration, and experimentation to address complex challenges (Cross, 2011).

At the outset of the workshop, LWHJ presented findings from the workshops with caregivers and professionals and a matrix analysis (see data analysis section), along with quotes illustrating the challenges encountered by caregivers and professionals. This aimed to provide software developers with a deeper understanding of the end users’ needs, context and the specific difficulties they face. Based on this, “How might We” questions were created and served as a practical guide for the software developers in developing suggestions for the proposed challenges (Siemon, 2018). The group brainstormed collaboratively to identify ideas for adaptions of the existing design of the messenger-like tool and how it could be used. The workshop lasted approximately 3,5 hours.

### 2.5 Data analysis

Data analysis was conducted in three overall phases as illustrated in Figure 3.

**Figure 3.**
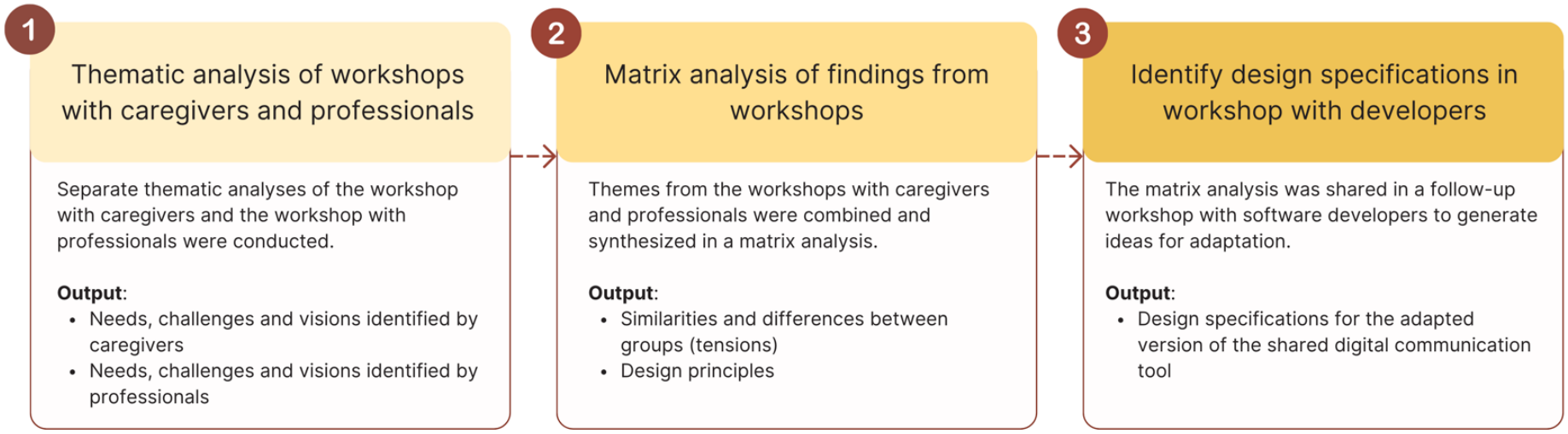
Overview of the data analysis process leading to the adaptation of the shared digital communication intervention.

#### Phase 1: Thematic analysis of workshops with caregivers and professionals

The workshops with caregivers and professionals were transcribed verbatim og analysed separately using inductive thematic analysis guided by Kvale & Brinkmann to identify common themes and insights from each group (Brinkmann and Kvale, 2015). Field notes and artifacts created during the workshops were also collected and coded. This was for example, notes written by caregivers and professionals about problems they have encountered related to communication and coordination, and ideas for adaptation of the intervention. Coding was conducted in Nvivo (QSR International, version 1.7.2) and following three overall steps (Appendix 4).

#### Phase 2: Matrix analysis

After the separated analyses of the workshops with caregivers and professionals, themes were combined and synthesized in a matrix analysis (Averill, 2002). This was a complementary analytic strategy to identify, display and interpret similarities and differences between the experiences and visions for the shared digital communication intervention as expressed by caregivers and professionals, respectively (Averill, 2002). Codes and themes from the thematic analysis was entered into a structured table, alongside relevant quotations from caregivers and professionals, and in this way the matrix enabled mapping and comparison (Averill, 2002). On one hand, similarities in themes from the workshop with caregivers and the workshop with professionals could highlight key steps in the subsequent design process (adaptations that are definitely needed), and on the other hand differences in themes between the groups could reveal tensions that were crucial to consider when adapting the intervention to the context. This approach ultimately helped identify and generate design principles (Fu et al., 2016, 2015).

#### Phase 3: Translation of findings into design specifications

The design principles generated through the matrix analysis served as initial design requirements during the subsequent workshop with the software developers. The developers translated these requirements into concrete design specifications while considering technical feasibility, platform architecture, and user workflows. In this way, the workshops with caregivers and professionals formed the basis of the development process with the software developers. The workshop with software developers was transcribed using only summary transcription, capturing the main points and key discussions and analysed using a design-oriented directed content analysis (Assarroudi et al., 2018; Elo and Kyngäs, 2008).

### 2.6 Involvement of people with lived experiences

People with lived experience is actively involved in the SHARE IT project, where both caregivers and professionals are represented in the advisory board and in a user panel. They contribute to defining the overall direction and purpose of the project, and we engage in ongoing dialogue to ensure that their input maintains the relevance of the project to real-world integrated care challenges. This study was conducted as a co-design process in which caregivers and professionals identified needs, challenges, and generated ideas for adapting the shared digital communication intervention to their context. The workshops were highly participatory, with the majority of time devoted to group work and shared dialogue. We chose to conduct separate workshops for caregivers and professionals to create a safe space in which each group could express their respective challenges and needs. During the subsequent data analysis, we paid particular attention to ensuring that the needs and ideas of both groups were given equal weight. The findings from the workshops were also discussed at a subsequent advisory board meeting attended by both caregivers and professionals. In this way, the contributions from people with lived experience truly informed the adaptation of the intervention and its alignment with everyday life and practice.

## 3. Ethical Considerations

Upon inquiry to the local ethics committee (North Denmark Region), the study did not require ethical approval; however, it was registered at the Research Database of North Denmark Region with journal number: F2024-178. The participation of caregivers, professionals and software developers was voluntary and informed consent was obtained prior to the workshops. Data collection, storage and management was conducted in concordance with the General Data Protection Regulation (GDPR).

## 4. Results

### 4.1 Characteristics of co-designers

Characteristics of caregivers (n=8), professionals (n=14) and software developers (n=4) attending the three workshops are presented in Table 3. The workshop with caregivers was initially planned to include 12 caregivers, however; one caregiver canceled participation the day before the workshop, two caregivers (a couple) cancelled on the day of the workshop due to their child’s illness, and one caregiver did not show up. None of the caregivers who attended the workshop were caregivers of the same child, meaning that each caregiver contributed experiences from a unique case.

**Table 3.** Characteristics of co-designers in the three workshops.

| **Co-designers** |  |  |  |
|---|---|---|---|
| **C/P** | **Caregivers (workshop one)** | **Professionals (workshop two)** | **Software developers (workshop three)** |
| **1** | *Parent* (f) of a child with cerebral palsy; nonverbal, uses wheelchair. | *Nurse* (f) from hospital with 10 years of experience\*. | *Software developer* (m) with 13 years of experience in company. |
| **2** | *Parent* (m) of a child with a syndrome; heart disease and severe scoliosis. | *Physiotherapist* (f) from hospital with 23 years of experience. | *Multimedia designer* (m) with three years of experience in company. |
| **3** | *Parent* (f) of a child with a genetic condition; deaf, blind, organ anomaly, and physical impairment. | *Occupational therapist* (f) from hospital with 18 years of experience. | *Head of department and software developer* (m) with 23 years of experience in company. |
| **4** | *Parent* (f) of a child with congenital virus infection; severe brain damage, reduced mobility, and impaired speech. | *Social worker* (f) from the municipality with 15 years of experience. | *Account manager* (f) with five years of experience in company. |
| **5** | *Parent* (f) of a child with cerebral palsy; nonverbal, uses wheelchair. | *Social worker* (f) from the municipality with six years of experience. |  |
| **6** | *Parent* (m) of a child with a rare genetic disorder; delayed development, immune problems, etc. | *Physiotherapist and department manager* (f) in the Danish Educational-Psychological Counselling service (PPR) with 21 years of experience. |  |
| **7** | *Parent* (f) of a child with cerebral palsy and epilepsy; reduced mobility, vision and speech. | *Occupational therapist* (f) in the PPR with 13 years of experience. |  |
| **8** | *Parent* (m) of a child born with limb defect; reduced mobility | *Teacher* (f) at a preschool for children with special needs with seven years of experience. |  |
| **9** |  | *Educational leader* (f) at a preschool for children with special needs with 10 years of experience. |  |
| **10** |  | *Private practice physiotherapist* (m) with two years of experience. |  |
| **11** |  | *Private practice physiotherapist* (f) with six years of experience. |  |
| **12** |  | *Teacher* (f) at a school for children with special needs with four years of experience. |  |
| **13** |  | *Occupational therapist* (f) from municipality with 13 years of experience. |  |
| **14** |  | *Physiotherapist* (f) from municipality with four years of experience. |  |

### 4.2 Results of the data analysis from the workshop with caregivers

Analysis of data from the workshop with caregivers generated four overall themes and additional subthemes highlighted in Figure 4 and elaborated in a narrative description with quotes.

**Figure 4.**
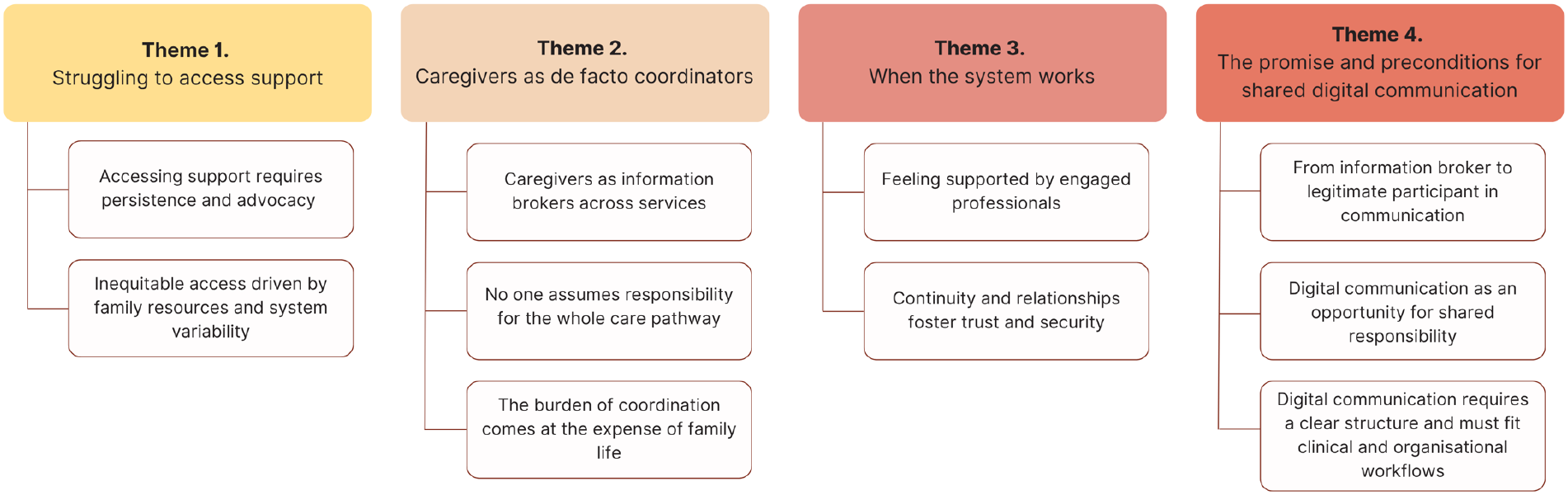
Themes and subthemes from analysis of data from the workshop with caregivers.

#### Theme 1: Struggling to access support

Caregivers experience that support is often dependent on the child’s needs being formally recognized by the system. This is particularly evident among caregivers of children with rare diseases, for whom a diagnosis serves as a gateway to accessing services, communication with relevant professionals and interventions.

> *‘When the child does not have a diagnosis, you are just left in an open box (…) You become a ping-pong ball being passed around, and it’s exhausting.’* (C4)

However, in everyday life when new assistive devices or specialized services from the municipality are needed, it is also described as an uphill battle, requiring caregivers to develop extensive knowledge of regulations, available services, and professional terminology. Caregivers feel they have to persistently argue their child’s case, provide documentation, and know what support to request. Moreover, support is not always experienced as stable, but can be reduced or removed, requiring families to repeatedly justify their needs.

Caregivers also describe considerable variation in access to support depending on municipal practices, individual professionals, and families’ own resources and networks. As such, caregivers with greater knowledge, economic opportunities, professional backgrounds, or supportive networks are perceived as better equipped to navigate the system, creating a sense of inequity in access to care and support.

> *‘And then having to fight to actually secure a place in a specialized institution for your child, because the municipality, which has never met him, decides that this is not what he needs. It becomes this endless struggle.’* (C3)

Caregivers’ experiences suggest that access to support is shaped not only by the child’s needs, but also by the individual family’s capacity to navigate a fragmented and complex system.

#### Theme 2: Caregivers as de facto care coordinators

Caregivers describe taking responsibility for maintaining an overview of their child’s needs across fragmented services. Due to insufficient information sharing between professionals, they repeatedly recount their child’s history, transfer documentation, and communicate relevant knowledge across organisational boundaries. Although caregivers recognize the need to take the lead and are willing to do so, they describe carrying a responsibility they have neither formally chosen nor been prepared for.

> *‘As parents, we need to know everything about our children, including the problems and challenges that we cannot necessarily describe using professional language.’* (C1)

Caregivers also highlight that no single professional or service consistently assumes responsibility for the child’s overall care pathway. Instead, services are experienced as organized around separate professional domains, while caregivers experience their child’s needs as interconnected and requiring a holistic approach. The involvement of multiple actors, unclear responsibilities, and limited communication across sectors contribute to caregivers’ perception that they have to create the necessary links between services themselves.

> *‘I think the issue is that I am the one responsible for passing on the information, and also for physically sending the documentation.’* (C5)

This coordination role has substantial consequences for family life. Caregivers describe the practical and emotional demands of managing communication, appointments, documentation, and follow-up activities alongside their everyday responsibilities as caregivers. The time and energy spent navigating the system reduces their opportunity to focus on being present with their child and family and contributes to feelings of exhaustion and frustration.

> *‘You spend so much time on all of this, and that time has to come from somewhere else - from your other children, for example (…) I had more than a year where I was not working. The amount of time I spent managing everything related to our son was enormous. There was always something happening: first a surgery, and then he developed epilepsy and so on. The amount of time spent on communication and coordination would never would have been possible if I had been working full time.’* (C7)

Several caregivers express a profound longing to simply be able to focus on being a parent.

Thus, caregivers’ experiences suggest that the absence of integrated coordination mechanisms results in a transfer of organisational work to families, giving caregivers responsibility for maintaining continuity across a fragmented system.

#### Theme 3: When the system works

Despite describing a fragmented system, caregivers also highlight how committed professionals can create a sense of coherence and support. Professionals who understand the child and family, recognize the child beyond their diagnosis, and actively seek solutions provide reassurance and reduce caregivers’ sense of having to navigate the system alone. One caregiver describes the difference a particular doctor has made:

> *‘He has relieved a huge burden for us, particularly in relation to our daughter’s condition. He has been curious, he has supported us, and whenever challenges have arisen, he has been there. He is the first person we have encountered within the system who has been like that.’* (C4)

Continuity in professional relationships is also important for building trust and security. Caregivers value professionals who know their child’s history and individual needs, sparing them from repeatedly explaining their situation. Conversely, frequent changes in professionals can lead to loss of knowledge, uncertainty, and the need to rebuild trust. When communication and coordination work well, caregivers often describe this as a matter of ‘luck’, suggesting that such support is incidental rather than a reliable feature of the system.

> *‘We have been fortunate that the social worker we were assigned when she was X years old is still our social worker today. That is quite extraordinary; it is not the norm. It has made a huge difference.’* (C3)

These experiences suggest that integrated care is not experienced solely as a result of organisational structures or formal coordination mechanisms, but is often dependent on individual professionals who take responsibility and demonstrate commitment.

#### Theme 4: The promise and preconditions for shared digital communication

Caregivers view shared digital communication as a potential way to reduce the burden of information management and improve cross-sector coordination. However, its value depends on whether it addresses existing challenges related to coordination, information sharing and responsibility.

> *‘I can think of a specific situation in our family…(where it would have been helpful). Our daughter had just started school, and the hospital recommended that she should attend school for a certain number of hours, but the municipality questioned that recommendation. In that situation, it would have been incredibly helpful. It would have made so much sense because I would not have had to sit there and ‘play doctor’ when talking to the municipality. That is where I can really see this working.’* (C6)

Caregivers describe that direct communication between professionals and the sharing of videos, plans, and documents may reduce caregivers’ role as information brokers, support their participation in care, and, in som cases, potentially avoid unnecessary appointments.

> *‘Going to an appointment is always an all-day project for us. The only thing they wanted to assess was how he was walking. I could have recorded a video and sent it to them.’* (C7)

However, caregivers’ express concerns that the intervention might place additional demands on professionals and question whether professionals will have the capacity and willingness to engage. They therefore emphasize the need for clear roles, responsibilities, and expectations to prevent additional demands and avoid the platform becoming an open consultation service.

> *‘If, for example, a doctor suddenly has all their patients (on this platform), who may normally only be seen twice a year, starting to send pictures several times a month and expecting a response, then there needs to be clear expectation-setting with parents. This should not become an open consultation where parents can ask information every time, they are uncertain.’* (C6)

Caregivers also want dialogues to focus on clearly defined tasks, with influence over their purpose and participants. This reflects concerns that information shared across professional contexts may be interpreted differently and have unintended consequences.

> *‘I would like to have a say in defining the topic, or at least ensure that it is clearly delimited. I have experienced several times that doctors, with the best intentions, write something in the medical record and forward it to the municipality to help us obtain a support aid or resolve a specific problem. However, the outcome can be the opposite of what was intended, because the information is interpreted differently (…).’* (C2)

Finally, caregivers emphasize that written dialogues should be structured and have a clear endpoint. Overall, shared digital communication is considered promising, provided that it reduces rather than shifts the coordination burden onto families.

### 4.3 Results of the data analysis from the workshop with professionals

Analysis of data from the workshop with professionals produced five overall themes and additional subthemes highlighted in Figure 5 and further elaborated in a narrative description with quotes from professionals.

**Figure 5.**
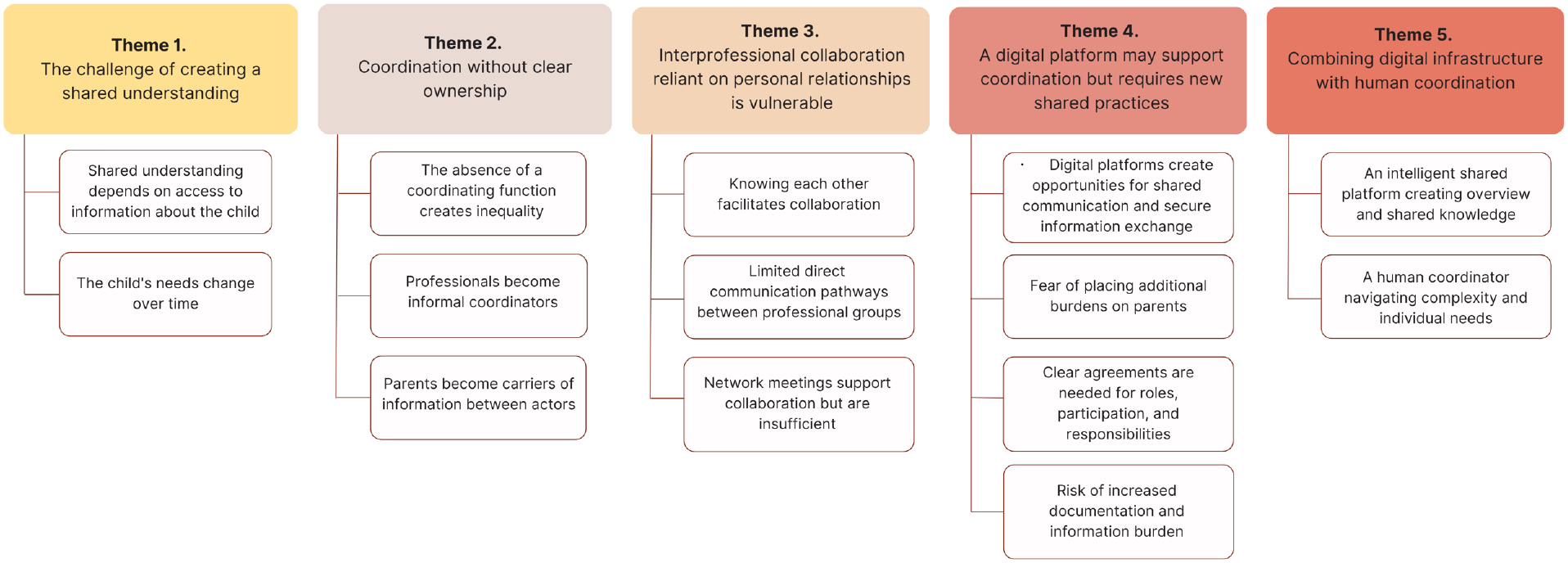
Themes and subthemes from analysis of data from the workshop with professionals.

#### Theme 1: The challenge of creating a shared understanding

Professionals emphasize that supporting children with physical disabilities requires a holistic understanding of the child’s abilities, development, treatment goals, and everyday needs. However, this is difficult to achieve when knowledge is distributed across professionals, organisations, and information systems, creating uncertainty about how best to support the child.

> *‘You can easily feel uncertain and start questioning whether you are doing something that could be harmful (when you lack information). Is there something I should avoid doing? When is he allowed to do that and that (after surgery)?’* (P6)

Limited access to shared information also means that professionals rely on caregivers to relay information, hindering timely and coordinated support as the child’s needs evolve. Furthermore, professionals may have different priorities based on their professional perspectives. Without a shared overarching goal, these differences can leave caregivers responsible for navigating competing perspectives and maintaining an overview of the child’s needs.

> *‘There are so many (professionals) involved, and so many professionals who genuinely want the best for the child and the family. I recognize that this can be a positive thing, but it also becomes problematic because they represent different views on what is important for the child and family (…) and the family somehow has to navigate these. We lack an overall overview.’* (P7)

These findings highlight the need for structures that enable shared understanding and alignment across professional and organisational boundaries.

#### Theme 2: Coordination without clear ownership

Professionals describe coordination across professionals, sectors, and services as lacking clear responsibility. Although individual actors coordinate within their own areas, no one has overall responsibility for ensuring continuity across the child’s care pathway. Consequently, professionals who have frequent contact with families or are easily accessible often assume an informal coordinating role by connecting actors, following up, and sharing information.

> *‘In the specialized preschool setting, we often take on a coordinating role, although it is not formally defined or explicitly assigned. This becomes particularly evident when children transition into the school system. We try to establish a network around the child and support families in identifying where to seek help, and for some families, we actively assist in making those contacts.’* (P9)

From the perspective of private providers, additional barriers to communication and coordination arise from the lack of financial incentives to participate in cross-sector collaboration.

> *‘Many forms of communication do not fit into the existing economic framework, such as informal conversations, text messages, or phone calls from parents. I experience this as a significant challenge. As a result, I sometimes limit my communication with parents, keeping it to a need-to-know basis or the absolute minimum required to ensure an appropriate course of care with the information available to me. I rely on parents to provide the information I need, for example during consultations, rather than having to seek it out myself outside of working hours.’* (P10)

As a result, professionals and caregivers often fill coordination gaps by transferring information themselves, placing an additional burden on families. Professionals also point to a growing private market for social workers, which may widen inequalities as access to such support depends on families’ financial resources.

#### Theme 3: Interprofessional collaboration reliant on personal relationships is vulnerable

Professionals describe established relationships between colleagues, including knowledge of each other’s roles and contact pathways, as important facilitators of interprofessional collaboration. However, access to these informal networks may depend on professional experience and personal connections, leaving newer professionals at a disadvantage.

> *‘I have developed certain lifelines within the system. I know that I can contact him or her if needed. However, this also makes the process somewhat dependent on chance, because it relies on the people I happen to know.’* (P4)

Although formal network meetings support shared understanding, professionals consider them insufficient due to their infrequency and limited ability to support ongoing communication. They may also be experienced by caregivers as overwhelming or insufficiently informative and inclusive.

Professionals further point to existing coordination structures and projects that are not available to all children. For example, children with acquired brain injuries or cancer may have access to dedicated cross-sector coordination, unlike most children with physical disabilities.

> *‘In these cases, parents know that, until their child turns 18, there is a brain injury centre that has a coordinating role and can be contacted for support. However, this only applies to a small group of children, and one could argue that a similar structure would be equally relevant for many other groups of children.’* (P2)

Overall, interpersonal relationships are an important source of coherence, but reliance on individual networks makes collaboration vulnerable to staff turnover and organisational differences. Existing coordination structures in other care pathways may offer valuable lessons for improving support for a broader group of children.

#### Theme 4: A digital platform may support coordination but requires new shared practices

Professionals see a shared digital platform as an opportunity to improve coordination through task-specific communication, information sharing, and follow-up across caregivers and professional and organisational boundaries. They consider it particularly relevant during transitions and when introducing new interventions.

> *‘I think it can be useful in relation to more clearly defined and specific interventions or pathways (…) Because these are very much centered around the physical aspects. And beyond this, it may also be relevant during transitions, such as when a child is referred to a specialized educational setting or moves from preschool to school, points where multiple actors become involved.’* (P8)

However, professionals emphasize that a platform alone cannot resolve coordination challenges. Clear agreements are needed regarding who participates, what information is shared, who responds, and within what timeframe. Without these, increased transparency may add to caregivers’ responsibility for monitoring communication rather than relieve their burden.

> *‘We talked about the pros and cons of parents being able to read along with what is being written between professionals, because we also have to be mindful of them. If it was intended as a way of relieving some of their burden, by allowing us to use it as a means of communication, but then they suddenly find themselves thinking, “Oh, now they (professionals) have written to each other again, and now I have to keep track of what they have written.” Then we might actually end up placing an additional burden on them.’* (P1)

Professionals also anticipate that easier access to communication may increase their workload. They therefore stress the need to allocate dedicated time for platform-related communication, referred to as ‘app time’.

Finally, information sharing raises concerns about privacy, consent, and GDPR, particularly as repeated parental consent is often required before professionals can communicate or share information. Overall, professionals emphasize the need to balance accessibility and transparency with appropriate boundaries for sensitive information, supported by shared communication practices and organisational agreements.

#### Theme 5: Combining digital infrastructure and human coordination

Professionals highlight the need to combine technological support with human coordination. In the workshops, they developed the concepts of ‘Preben’ and ‘Sonja’ (Danish names) to represent two complementary forms of support: an automated digital overview and a human coordinating function.

’Preben’ represents a vision of an intelligent, shared platform providing an updated overview of the child’s situation, involved professionals, ongoing activities, and relevant information. Through secure, consent-based information sharing, such a system could facilitate access to information, reduce duplicate documentation, and ensure continuity across transitions and organisational boundaries.

> *‘The idea was that “Preben” would collect data from different sources and create a shared overview. For example, if a rehabilitation plan for hand training was created for a child, professionals could access information about the hand, including previous assessments, clinical notes, and relevant documentation. This would provide a basis for deciding the next steps.. (…). Parents could also be included in this overview, allowing professionals to consider the wider context of the child’s life, including siblings, school experiences, and potential needs for additional support.’* (P3)

However, professionals emphasize that technology alone cannot fulfill the coordinating role. ‘Sonja’ represents a human coordinator who ensures that relevant professionals are involved and addresses the relational and emotional aspects that cannot be captured through digital information alone.

> *’So, in order to prevent the parents from ending up being the ones who take on the coordinating role after all, “Sonja” steps in and takes on that role instead.’ (P1)*

Thus, professionals see effective coordination as requiring both shared digital infrastructure and human responsibility.

### 4.3 Matrix analysis

The matrix analysis identified key tensions and opportunities related to the shared digital communication intervention. Across the workshops with caregivers and professionals, both groups identified fragmented coordination and insufficient information sharing as central challenges, while responsibility for managing these challenges was often placed on caregivers and individual professionals rather than being structurally embedded within the care system. Caregivers described the burden of collecting, interpreting, and transferring information across services, while professionals viewed this as a consequence of lacking formal coordination structures.

A related finding concerned the need for a shared understanding of the child across professional and organisational boundaries. Caregivers emphasized the importance of professionals having access to the child’s history, needs, and everyday circumstances without requiring families to repeatedly provide this information. Professionals similarly described difficulties in developing a holistic understanding when knowledge is distributed across multiple actors and systems. The matrix thus identified a shared need for a common overview to support continuity and alignment, while highlighting that shared knowledge requires not only access to information but also agreement on its relevance, interpretation, and associated responsibilities.

The matrix analysis also revealed tensions regarding the potential role of digital communication. Both groups saw opportunities for improved information exchange, continuity, transparency, and reduced repetition, but expressed concerns about creating additional demands. Caregivers worried about overburdening professionals, while professionals worried about overburdening caregivers. Moreover, they were concerned that shared digital communication could add to documentation and communication demands and create uncertainty about responsibilities and response expectations. Finally, both groups emphasized that digital infrastructure alone cannot resolve the complexity of coordinating support. Caregivers highlighted the importance of professionals who know the child and family, while professionals emphasized the need for a coordinating function that ensures that relevant actors are involved.

The matrix analysis identified several design principles for a shared digital communication intervention including 1) a shared child overview, 2) shared communication with transparent roles and responsibilities, 3) to ensure equitable access to support, 4) to support continuity across professionals, 5) structured communication divided in specific tasks, 6) role-based access and consent management (caregiver control), 7) to use the tool as transitions support and, 8) to combine the digital platform with human coordinator support (Appendix 5).

### 4.4 Translation of findings into design specifications through workshop with software developers

The workshop with software developers translated the needs and tensions identified among caregivers and professionals into design specifications, ensuring alignment with clinical practice, the existing messenger-like platform, and its future development possibilities. This involved prioritizing features and making compromises.

For example, the analysis identified a need for a more comprehensive cross-system overview of the child, corresponding to the ‘Preben’ concept developed by professionals. However, as the messenger-like tool is primarily a secure communication platform rather than an integrated clinical information system, this was considered beyond the scope of the current intervention. The adapted intervention therefore focused on shared communication and coordination within the existing system, while cross-system data integration was retained as a potential direction for future development.

Furthermore, based on the design principles of ‘shared communication with transparent roles and responsibilities’ and ‘structured communication divided into specific tasks’, a distinction was made between primary professionals and professionals with sporadic or one-off involvement. This distinction highlighted the need for a shared coordination dialogue involving primary professionals who have ongoing and repeated contact with the child or caregivers, such as physicians, physiotherapists, social workers, and teachers. In addition, task-specific dialogues were identified as necessary to accommodate structured communication divided into specific tasks and to be able to include professionals with more sporadic involvement.

Finally, the integration of a messenger-like tool with a human digital coordinator was further developed and specified. Establishing a professionally qualified coordinator role was beyond the scope of this project, as such a function would be resource-intensive and raise questions regarding which organisation or sector, regional, municipal, or private providers should assume responsibility for the role. Instead, a human digital coordinator role was proposed as a feasible solution within the current project. The human digital coordinator could be a project staff member responsible for ensuring that relevant professionals were included in the communication, based on caregivers’ preferences, establish and close dialogues and clarify expectations regarding platform use, particularly during the initial phase.

Overall, the identified design specifications covered the following areas related to content and context: intervention content and functionality, conditions for access, delivery, coordination, and organisational support (Appendix 6).

## 5. The adapted shared digital communication intervention

The adapted intervention comprises five interrelated elements to support shared digital communication (Figure 6). Together, these elements are designed to support shared knowledge, distribute coordination responsibilities, and structure communication around the child’s specific needs and processes.

**Figure 6.**
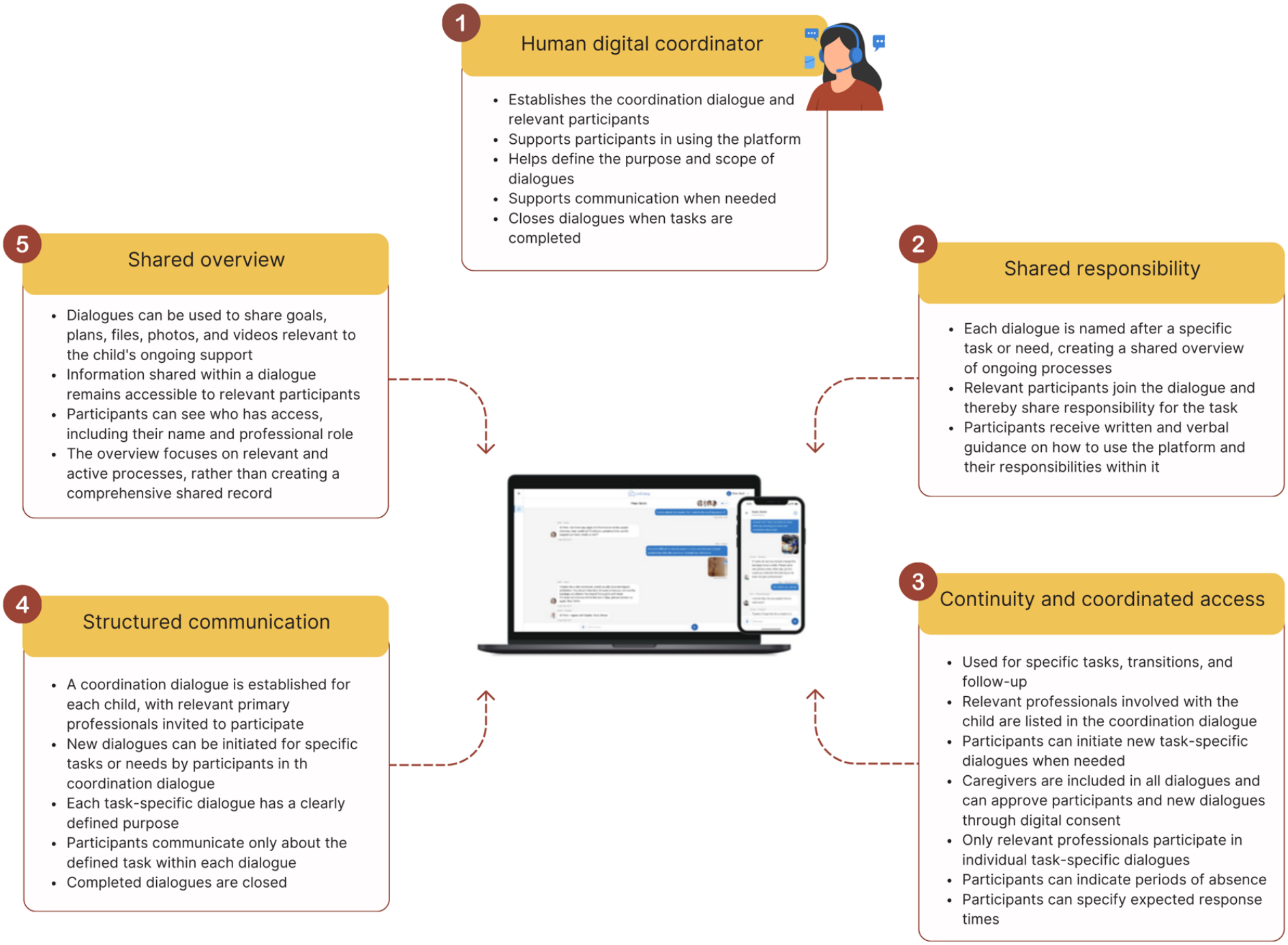
An overview of the adapted intervention for shared digital communication between caregivers of children with physical disabilities and professionals across disciplines and sectors.

## 6. Discussion

### 6.1 Primary results and comparison to previous literature

This study explored how an existing shared digital communication intervention could be adapted to support communication and coordination between caregivers of children with physical disabilities and professionals across sectors. Both groups described fragmented coordination and distributed knowledge, leaving caregivers as de facto coordinators of their child’s care. Experiences of integrated care were closely linked to whether someone took responsibility for creating coherence across the care pathway. Although shared digital communication was seen as a way to redistribute this responsibility, it could also create additional workload. The adapted intervention therefore incorporated shared responsibility, continuity, structured communication, a shared overview, and a human digital coordinating function.

Responsibility for coordination and continuity

A central finding was that caregivers frequently assumed responsibility for coordinating care by collecting and transferring information, interpreting professional assessments, following up on decisions, and connecting professionals. Professionals similarly described relying on caregivers to transfer information between services. Thus, caregivers became an informal mechanism through which the system achieved continuity.

This finding relates to family-centred care, which positions caregivers as active partners and recognizes their knowledge of their child’s needs and everyday life life (King and Chiarello, 2014). However, our findings highlight an important distinction between caregiver participation and caregiver responsibility. Caregivers wanted to participate in communication and decisions but did not want to be responsible for coordinating fragmented services. Family-centred care should therefore not imply transferring more responsibility to families. Meaningful partnership requires professionals and organisations to provide structures that enable participation without making families compensate for system fragmentation.

This finding aligns with literature documenting the substantial coordination work undertaken by families of children with complex or long-term conditions (Gulmans et al., 2009). However, when integrated care depends on families’ ability to navigate professional and organizational boundaries, access may depend on family resources. Families with greater knowledge, financial resources, professional experience, or social networks may be better positioned to advocate for support (Bøttcher and Dammeyer, 2025), raising concerns about equity.

Professionals described a parallel challenge: coordination was valued but insufficiently anchored in formal responsibilities, leaving individual professionals to act as informal coordinators. This contrasted with caregivers’ accounts of when the system worked well, where professionals who knew the child, responded when needed, and took responsibility beyond their immediate roles provided relief, trust, and security. Together, these findings suggest that integrated care depends on someone taking responsibility for creating coherence across the care pathway.

This has implications for digital interventions. A communication tool can facilitate information sharing but cannot create integration if responsibility for interpreting information, initiating collaboration, and following up remains unclear. Technology-supported care depends on the interaction between technology, users, and organizational contexts (Greenhalgh et al., 2017). Shared digital communication should therefore support the relational and organizational work of coordination while reducing reliance on individual goodwill, personal networks, and families’ capacity to compensate for gaps in the system.

Digital communication and the distribution of responsibility

Caregivers and professionals highlighted a potential in direct communication and shared access to information, including documents, photographs, and videos, to reduce repetition and support collaboration, particularly during transitions and when several professionals work towards a shared goal. This aligns with literature suggesting the need for ongoing communication and collaboration between families and service providers (Almasri et al., 2018, 2011).

However, our findings reveal a potential paradox. Caregivers wanted greater access to professionals but were concerned about increasing their workload, while professionals welcomed better access to information but anticipated increased communication, documentation, and expectations of responsiveness. Thus, an intervention intended to reduce the burden could instead redistribute it. This reflects evidence that health technologies may create additional workload and require changes to existing workflows if not adequately integrated into organizational practice (Grossi et al., 2021).

The relevant question is therefore not simply whether communication can be made easier, but what communication is necessary, for whom, and who is responsible for acting on it. This informed the decision to structure communication around specific tasks, with clear objectives, defined participants, response expectations, and closure of dialogues.

### Digital infrastructure needs to be combined with human coordination

The co-design process highlighted that digital infrastructure needs to be supported by human coordination. Professionals’ conceptualisations of ‘Preben’ and ‘Sonja’ captured two complementary needs: a shared overview of relevant information and a coordinator who ensures that relevant actors are involved in meeting the family’s needs.

Although the comprehensive cross-system overview envisioned through ‘Preben’ was beyond the scope of this study, the underlying need informed the intervention adaptation. Similarly, allocating a dedicated care coordinator to all families, as envisioned through ‘Sonja’, was beyond the project’s scope due to the resources required. Instead, the concept was adapted into a human digital coordinator role. This was considered a feasible way to support shared digital communication within existing legal and organizational frameworks, while ensuring that communication remained purposeful and coordination did not default to the family.

The intervention thus combined digital and relational coordination where relevant professionals could participate in dialogues, caregivers could influence who was involved, and a human digital coordinator could establish and support the communication structure. This reflects literature conceptualising digital technologies as embedded within wider organizational and relational infrastructures (Greenhalgh et al., 2019). It also addresses the vulnerability of relying on personal relationships and knowing ‘the right person’. Although relational continuity is important, it also requires organizational structures and shared responsibility across professional boundaries (Ljungholm et al., 2022).

### The co-design process revealed tensions that shaped the adaption of the intervention

This study illustrates the value of involving different stakeholder groups in intervention adaptation. Although caregivers and professionals shared concerns about fragmentation and the lack of a holistic understanding of the child, their perspectives on the digital solution differed. Caregivers emphasized accessibility and transparency, whereas professionals highlighted workload, boundaries, and responsibility. The matrix analysis helped identify the interdependence of these perspectives rather than treating them as competing requirements.

This is particularly relevant to participatory approaches to integrated care, where co-design brings together different forms of knowledge and requires dialogue and negotiation across stakeholders (Masterson et al., 2024). Participation therefore did not simply generate desired functions but revealed tensions requiring design compromises, including balancing accessibility with workload and transparency with information boundaries. The adaptation consequently addressed not only the technology but also the practices and conditions in which it would operate. This reflects Stirman et al.’s (2013) conceptualisation of intervention adaptation, which encompasses changes to both intervention content and implementation context (Stirman et al., 2013). Further adaptation will be made based on a feasibility study.

### 6.2 Strengths and limitations

A key strength of this study is its participatory approach, which enabled caregivers’ and professionals’ experiential knowledge, alongside software developers’ practical considerations, to inform intervention adaptation from an early stage (Moore et al., 2021b). Early user involvement can improve the relevance and usability of digital health interventions by ensuring that solutions reflect users’ needs and contexts (Baines et al., 2022; Masterson et al., 2024). The matrix analysis also enabled examination of both convergence and tensions between stakeholder perspectives rather than treating either group as homogeneous (Averill, 2002).

The study also has limitations. Recruitment was primarily based on convenience sampling, which may have limited variation in perspectives and underrepresented experiences that more deliberate sampling could have identified (Malterud et al., 2016). Although convenience sampling may require a larger sample to achieve sufficient breadth, the specificity and diversity of experiences within an available sample cannot always be predicted (Malterud et al., 2016). Furthermore, organizational decision-makers were represented on the SHARE IT project’s advisory board, however their more direct involvement in the early adaptation process may have facilitated future implementation.

The caregiver group in the first workshop was relatively small (n = 8). However, the discussions generated rich and diverse insights, and groups of eight to 16 participants are considered appropriate for facilitating dialogue and co-constructing knowledge (Kanstrup and Bertelsen, 2011). No physicians participated in the professional workshop, although professionals working closely with physicians provided insights into their communication workflows. Nevertheless, this limits the breadth of professional perspectives represented.

## 7. Conclusion

This study suggests that challenges in communication and coordination for children with physical disabilities are closely related to a lack of structures supporting shared knowledge, continuity, and responsibility across organisational boundaries. A shared digital communication intervention should be embedded within clear roles, shared responsibility and human coordination. Further research is needed to assess its feasibility, acceptability, impact on caregivers and professional workload, and organizational implications.

## Data Availability

All data produced in the present study are available upon reasonable request to the authors

## Acknowledgements

We are grateful to the caregivers, professionals, and software developers who participated in the workshops and contributed their experiences, perspectives, and expertise to the co-design and adaptation of the intervention. We would also like to acknowledge Rikke Emilie Kildahl Lauritsen for her contribution to the data analysis process.

## Appendix 1a: GUIDED reporting guideline

### GUIDED – a guideline for reporting for intervention development studies

**Supplementary File 1:** Blank Checklist.

| Item description | Explanation | Page in manuscript where item is located | Other* |
|---|---|---|---|
| 1. Report the context for which the intervention was developed. | Understanding the context in which an intervention was developed informs readers about the suitability and transferability of the intervention to the context in which they are considering evaluating, adapting or using the intervention. Context here can include place, organisational and wider sociopolitical factors that may influence the development and/or delivery of the intervention (15). | Page 1-2 |  |
| 2. Report the purpose of the intervention development process. | Clearly describing the purpose of the intervention specifies what it sets out to achieve. The purpose may be informed by research priorities, for example those identified in systematic reviews, evidence gaps set out in practice guidance such as The National Institute for Health and Care Excellence or specific prioritisation exercises such as those undertaken with patients and practitioners through the James Lind Alliance. | Page 1-2 |  |
| 3. Report the target population for the intervention development process. | The target population is the population that will potentially benefit from the intervention – this may include patients, clinicians, and/or members of the public. If the target population is clearly described then readers will be able to understand the relevance of the intervention to their own research or practice. Health inequalities, gender and ethnicity are features of the target population that may be relevant to intervention development processes. | Page 2 |  |
| 4. Report how any published intervention development approach contributed to the development process | Many formal intervention development approaches exist and are used to guide the intervention development process (e.g. 6Squid (16) or The Person Based Approach to Intervention Development (17)). Where a formal intervention development approach is used, it is helpful to describe the process that was followed, including any deviations. More general approaches to intervention development also exist and have been categorised as follows (3):- Target Population-centred intervention development; evidence and theory-based intervention development; partnership intervention development; implementation-based intervention development; efficacy-based intervention development; step or phased-based intervention development; and intervention-specific intervention development (3). These approaches do not always have specific guidance that describe their use. Nevertheless, it is helpful to give a rich description of how any published approach was operationalised | Page 1 |  |
| 5. Report how evidence from different sources informed the intervention development process. | Intervention development is often based on published evidence and/or primary data that has been collected to inform the intervention development process. It is useful to describe and reference all forms of evidence and data that have informed the development of the intervention because evidence bases can change rapidly, and to explain the manner in which the evidence and/or data was used. Understanding what evidence was and was not available at the time of intervention development can help readers to assess transferability to their current situation. | Page 1-2 |  |
| 6. Report how/if published theory informed the intervention development process. | Reporting whether and how theory informed the intervention development process aids the reader’s understanding of the theoretical rationale that underpins the intervention. Though not mentioned in the e-Delphi or consensus meeting, it became increasingly apparent through the development of our guidance that this theory item could relate to either existing published theory or programme theory | Page 1 |  |
| 7. Report any use of components from an existing intervention in the current intervention development process. | Some interventions are developed with components that have been adopted from existing interventions. Clearly identifying components that have been adopted or adapted and acknowledging their original source helps the reader to understand and distinguish between the novel and adopted components of the new intervention. | Page 1 + Appendix 6 |  |
| 8. Report any guiding principles, people or factors that were prioritised when making decisions during the intervention development process. | Reporting any guiding principles that governed the development of the application helps the reader to understand the authors’ reasoning behind the decisions that were made. These could include the examples of particular populations who views are being considered when designing the intervention, the modality that is viewed as being most appropriate, design features considered important for the target population, or the potential for the intervention to be scaled up. | Page 6-12 |  |
| 9. Report how Potential stakeholders can include patient and community representatives, stakeholders local and national policy makers, health care providers and those paying for or contributed to the commissioning health care. Each of these groups may influence the intervention development process in different ways. Specifying how differing development process. groups of stakeholders contributed to the intervention development process helps the reader to understand how stakeholders were involved and the degree of influence they had on the overall process. Further detail on how to integrate stakeholder contributions within intervention reporting are available (19). |  | Page 2 + 4 |  |
| 10. Report how the intervention changed in content and format from the start of the intervention development process. | Intervention development is frequently an iterative process. The conclusion of the initial phase of intervention development does not necessarily mean that all uncertainties have been addressed. It is helpful to list remaining uncertainties such as the intervention intensity, mode of delivery, materials, procedures, or type of location that the intervention is most suitable for. This can guide other researchers to potential future areas of research and practitioners about uncertainties relevant to their healthcare context. | Appendix 6 |  |
| 11. Report any changes to interventions required or likely to be required for subgroups. | Specifying any changes that the intervention development team perceive are required for the intervention to be delivered or tailored to specific sub groups enables readers to understand the applicability of the intervention to their target population or context. These changes could include changes to personnel delivering the intervention, to the content of the intervention, or to the mode of delivery of the intervention. | - |  |
| 12. Report important uncertainties at the end of the intervention development process. | Intervention development is frequently an iterative process. The conclusion of the initial phase of intervention development does not necessarily mean that all uncertainties have been addressed. It is helpful to list remaining uncertainties such as the intervention intensity, mode of delivery, materials, procedures, or type of location that the intervention is most suitable for. This can guide other researchers to potential future areas of research and practitioners about uncertainties relevant to their healthcare context. | Page 15 |  |
| 13. Follow TIDieR guidance when describing the developed intervention. | Interventions have been poorly reported for a number of years. In response to this, internationally recognized guidance has been published to support the high-quality reporting of health care? interventions<sup>5</sup>and public health interventions<sup>14</sup>. This guidance should therefore be followed when describing a developed intervention. | We followed The FRAME instead because this was an adaptation (see Appendix 1b) |  |
| 14. Report the intervention development process in an open access format. | Unless reports of intervention development are available people considering using an intervention cannot understand the process that was undertaken and make a judgement about its appropriateness to their context. It also limits cumulative learning about intervention development methodology and observed consequences at later evaluation, translation and implementation stages. Reporting intervention development in an open access (Gold or Green) publishing format increases the accessibility and visibility of intervention development research and makes it more likely to be read and used. Potential platforms for open access publication of intervention development include open access journal publications, freely accessible funder reports or a study web-page that details the intervention development process. | Yes |  |

\*e.g. if item is reported elsewhere, then the location of this information can be stated here.

---

**Appendix 1b:** Reporting adaptations using the FRAME.

| **FRAME dimension** | **Reporting category** | **Description of adaptation in this study** |
|---|---|---|
| 1. When and how | Timing and process | The modifications were made during the adaptation phase preceding feasibility testing, evaluation and implementation, as part of an iterative co-design process. This process involved three workshops with family caregivers, professionals from various sectors, and software developers. During these workshops, participants explored experiences and challenges related to communication and coordination, identified needs and design principles, and translated these into specifications for the adapted intervention. The research team facilitated the process and supported the incorporation of input from the involved parties into the adapted intervention. |
| 2. Planned or unplanned | Planned / proactive or unplanned / reactive | The modifications were planned and proactive. They were developed through a structured co-design process before the adapted intervention was implemented for feasibility testing in the new context. The adaptations were therefore not reactive responses to unforeseen implementation barriers. |
| 3. Who determined the modification | Decision-makers and contributors | The modifications were informed by the perspectives and contributions of caregivers, professionals from different sectors and software developers. The research team facilitated the co-design process, analyzed and synthesized the input, and supported the translation of identified needs into intervention specifications. The adaptations thus resulted from a collaborative process. The study does not specify a single decision-maker with final authority over each individual modification. However, the advisory board was consulted during the adaptation process. |
| 4. What was modified | Content and components | The existing shared digital communication intervention was adapted to support coordination and communication between caregivers and relevant professionals involved in the support of children with physical disabilities. The adapted intervention comprises five interrelated core elements:<br>1. Human digital coordinator: A human coordinator establishes the coordination dialogue and identifies relevant participants, supports participants in using the platform, helps define the purpose and scope of dialogues, supports communication when needed, and closes dialogues when tasks have been completed.<br>2. Shared responsibility: Each dialogue is named after a specific task or need to create a shared overview of ongoing processes. Relevant participants join the dialogue and thereby share responsibility for the task. Participants receive written and verbal guidance on using the platform and their responsibilities within it.<br>3. Continuity and coordinated access: The platform is used for specific tasks, transitions and follow-up. Relevant professionals involved with the child are listed in the coordination dialogue, and participants can initiate new task-specific dialogues when needed. Caregivers are included in all dialogues and can approve participants and new dialogues through digital consent. Only professionals relevant to the individual task participate in each task-specific dialogue. Participants can also indicate periods of absence and specify expected response times.<br>4. Structured communication: A coordination dialogue is established for each child, with relevant primary professionals invited to participate. Participants can initiate additional dialogues for specific tasks or needs. Each task-specific dialogue has a clearly defined purpose, and communication is restricted to the task defined within that dialogue. Completed dialogues are closed.<br>5. Shared overview: Dialogues can be used to share goals, plans, files, photographs and videos relevant to the child's ongoing support. Information shared within a dialogue remains accessible to relevant participants, who can see who has access, including participants' names and professional roles. The overview focuses on relevant and active processes rather than creating a comprehensive shared record. |
| 5. Level of delivery | Level at which the modification was made | The modifications were made at the intervention-delivery level, with implications for coordination across organizational and professional boundaries. The adapted intervention was designed for use by caregivers and relevant professionals involved in a child's support across sectors. The modifications were not intended as individual adaptations for particular caregivers or professionals, but as features of the intervention's overall structure and delivery. |
| 6. Nature of modifications | Content-level and context-level modifications | The adaptations involved both changes to intervention content and changes to the way the intervention was intended to be delivered and used in its new context.<br>At the content level, the adapted intervention specified five interrelated elements: a human digital coordinator, shared responsibility, continuity and coordinated access, structured communication, and a shared overview. These elements introduced or specified functions and procedures for establishing, organizing, facilitating and closing task-specific dialogues, as well as sharing and accessing relevant information.<br>At the context and delivery level, the intervention was adapted to support collaboration between caregivers and professionals across different sectors. This included clarifying participants' roles and responsibilities, giving caregivers control over participation through digital consent, and structuring communication around specific tasks and relevant participants. The adaptations also addressed the need for human support alongside the digital platform. |
| 7. Fidelity consistency | Fidelity-consistent / fidelity-inconsistent / unknown | The adaptations were intended to retain the overall purpose of the original intervention as a shared digital communication tool while modifying its content and delivery to fit the needs and context of caregivers and professionals supporting children with physical disabilities. The study did not include a formal assessment of intervention fidelity or explicitly establish which components of the original intervention were considered essential to its effectiveness. Consequently, the extent to which the modifications were consistent with intervention fidelity cannot be determined conclusively. |
| 8. Reasons for modifications | Intent and contextual factors | The modifications aimed to improve the intervention's relevance, acceptability and feasibility in a context characterized by fragmented communication and coordination across health, social care and educational services. The co-design process identified challenges relating to information sharing, continuity, unclear responsibilities and the coordination work undertaken by caregivers and professionals.<br>The five elements were developed to address these challenges by supporting shared responsibility, making communication more structured and task-specific, facilitating continuity across tasks and transitions, and providing access to relevant information for those involved. The human digital coordinator was included to support participants and help ensure that dialogues were established, purposeful and closed when tasks were completed.<br>The adaptations also sought to address the risk that a shared digital communication platform could create additional communication demands without adequately supporting coordination. By defining the purpose and scope of dialogues, clarifying responsibilities, limiting participation to relevant individuals and specifying expectations regarding availability and response times, the adapted intervention aimed to support purposeful communication while taking account of the different roles and working conditions of caregivers and professionals.<br>The contextual factors informing the adaptations included the involvement of multiple professional groups and organizational settings, differences in professional responsibilities and workflows, the need to protect information and manage access, and the practical and technical conditions for using a shared digital platform. |

**Appendix 2:**
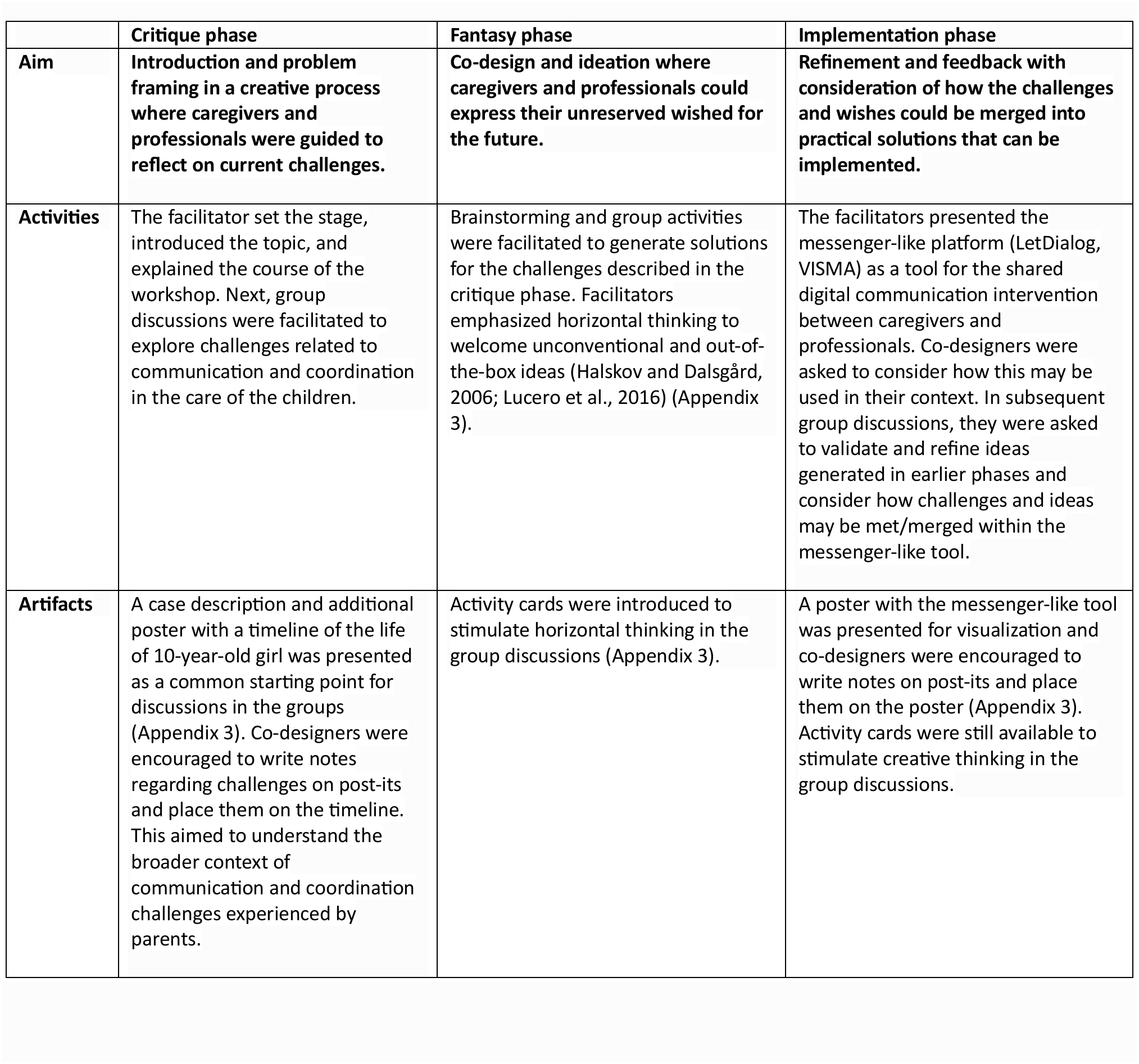
Summary of the action plan used for the workshops with caregivers and professionals.

## Appendix 3: Artifacts used during workshops with caregivers and professionals

Initially, a fictive case is presented to participants showing only the black text, creating the impression that the child had received adequate support and that her needs were met. After participants read the black text, the red text is introduced to provide additional context, facilitate a sense of understanding of all the challenges caregivers meet and framing the shared case for collaborative discussion in the three phases of the workshop.

### The case

Liv was born on March 18, 2015, at Rigshospitalet in Copenhagen. The birth was complicated, and Liv was diagnosed with cerebral palsy (CP) shortly after birth. After birth, Liv and her family spent ten weeks in the neonatal department. In the first years, Liv received a rehabilitation plan, but there were long waiting times for permanent therapists.

Applications for assistive devices and support were delayed due to bureaucracy and poor communication. Liv started in a special kindergarten, but resources were limited. The caregivers had to find external help themselves, which was time-consuming and expensive. Liv started in a public school with a special class, but the transition was chaotic due to lack of communication and coordination. The necessary assistive devices were not in place, and her support teachers were often replaced. The annual evaluations were marked by a lack of coordination and disagreements on goals and methods, which resulted in Liv’s needs not always being met.

The family had difficulty receiving adequate help at home and respite care, which led to the caregivers often feeling overwhelmed and burned out. There were few services tailored to Liv’s needs, and caregivers had to spend a lot of time and money finding relevant leisure activities. The family struggled to build a support network due to distance and waiting lists, which led to isolation for both Liv and the family.

By the age of ten, Liv is a brave and curious girl, but her development has been marked by many challenges within the system. Her motor skills could have been better with more effective physiotherapy. She thrives in school, but has experienced many changes and uncertainties, which have affected her learning and social development. The family continues to fight to receive the necessary support.

### Activity cards for workshop with caregivers and professionals

Activity cards aimed to represent visualizations within “stakeholders”, “system” and “technology” to promote horizontal thinking during the workshops fantasy phase. Each picture or graphic was printed individually on separate cards and placed at each table (group) and used by facilitators to boost creativity with participants.

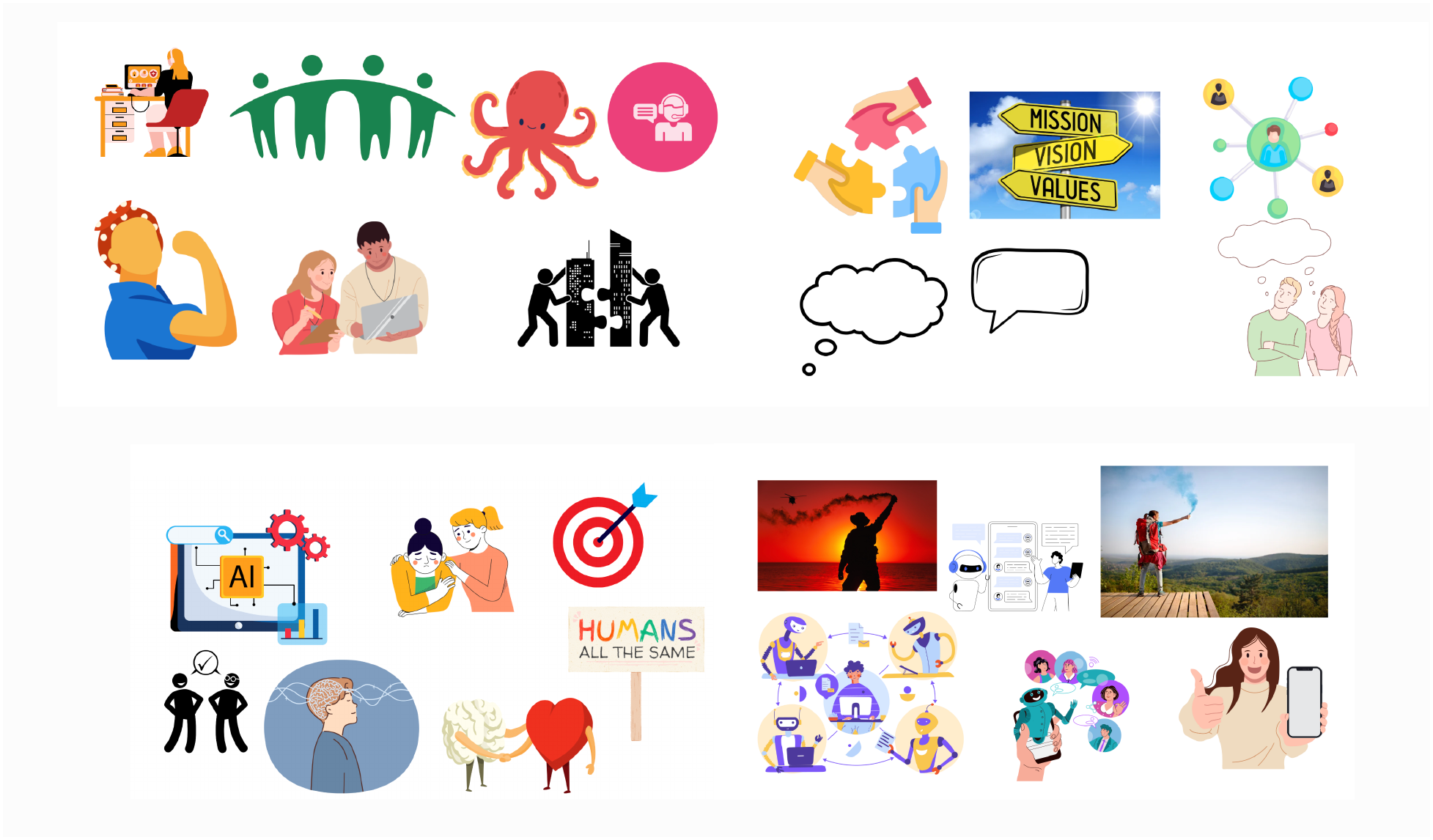
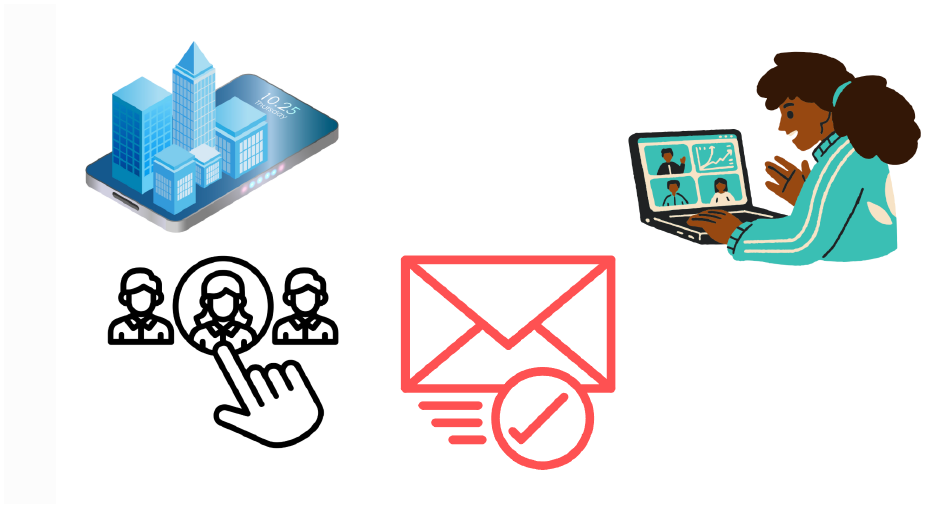

### Poster used in critique and fantasy phases

The poster was used during the critique and fantasy phases of the workshops with caregivers and professionals. Participants were asked to write or illustrate challenges (critique phase) and potential solutions (visions phase), placing them along Liv’s life timeline. This aimed to pinpoint critical moments where challenges emerged and identify opportunities for intervention based on participants’ insights.

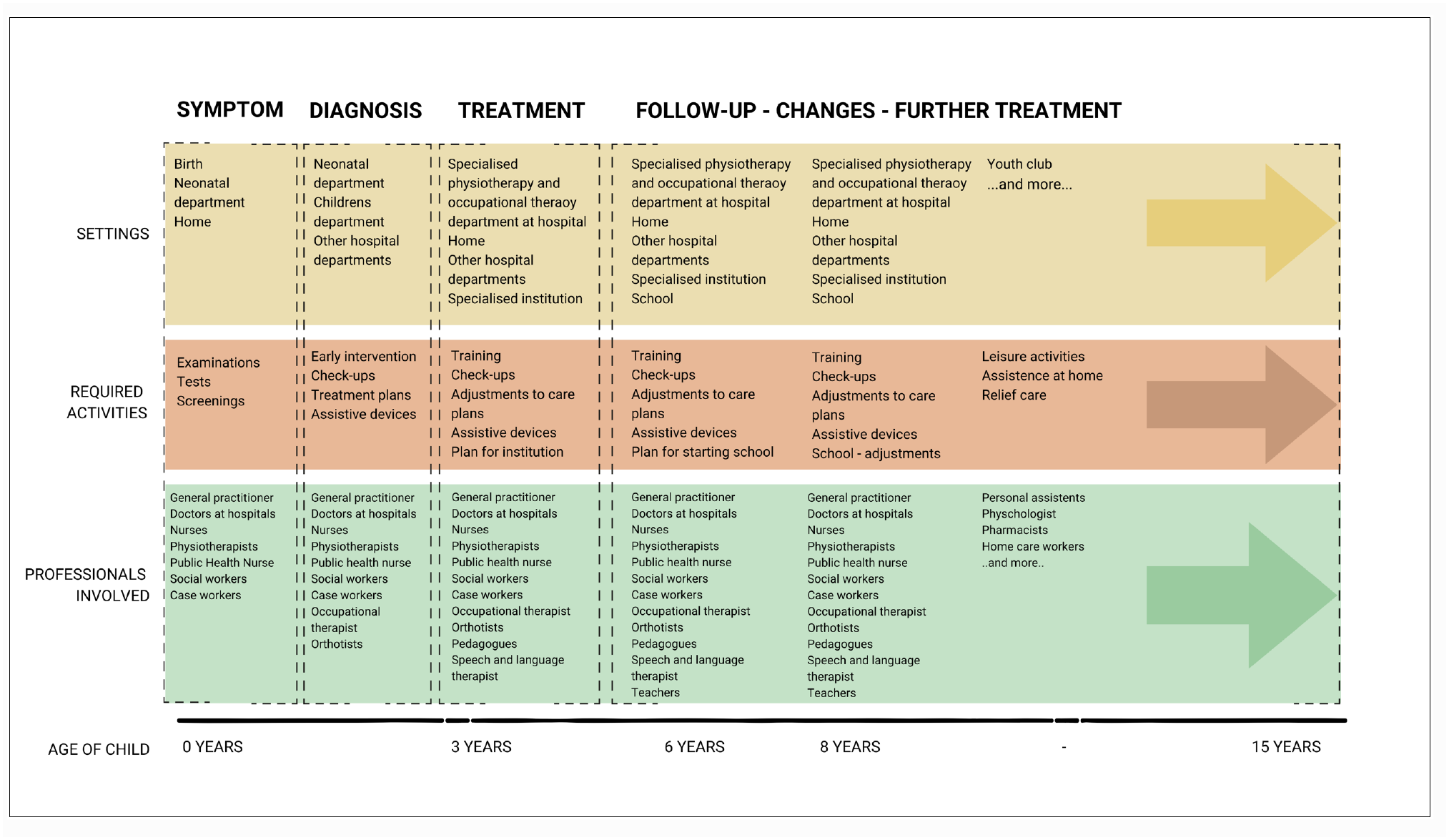

### Poster used in implementation phase

The poster was used during the implementation phase of the workshops with caregivers and professionals. This aimed to generate concrete ideas for adapting the tool to the new context.

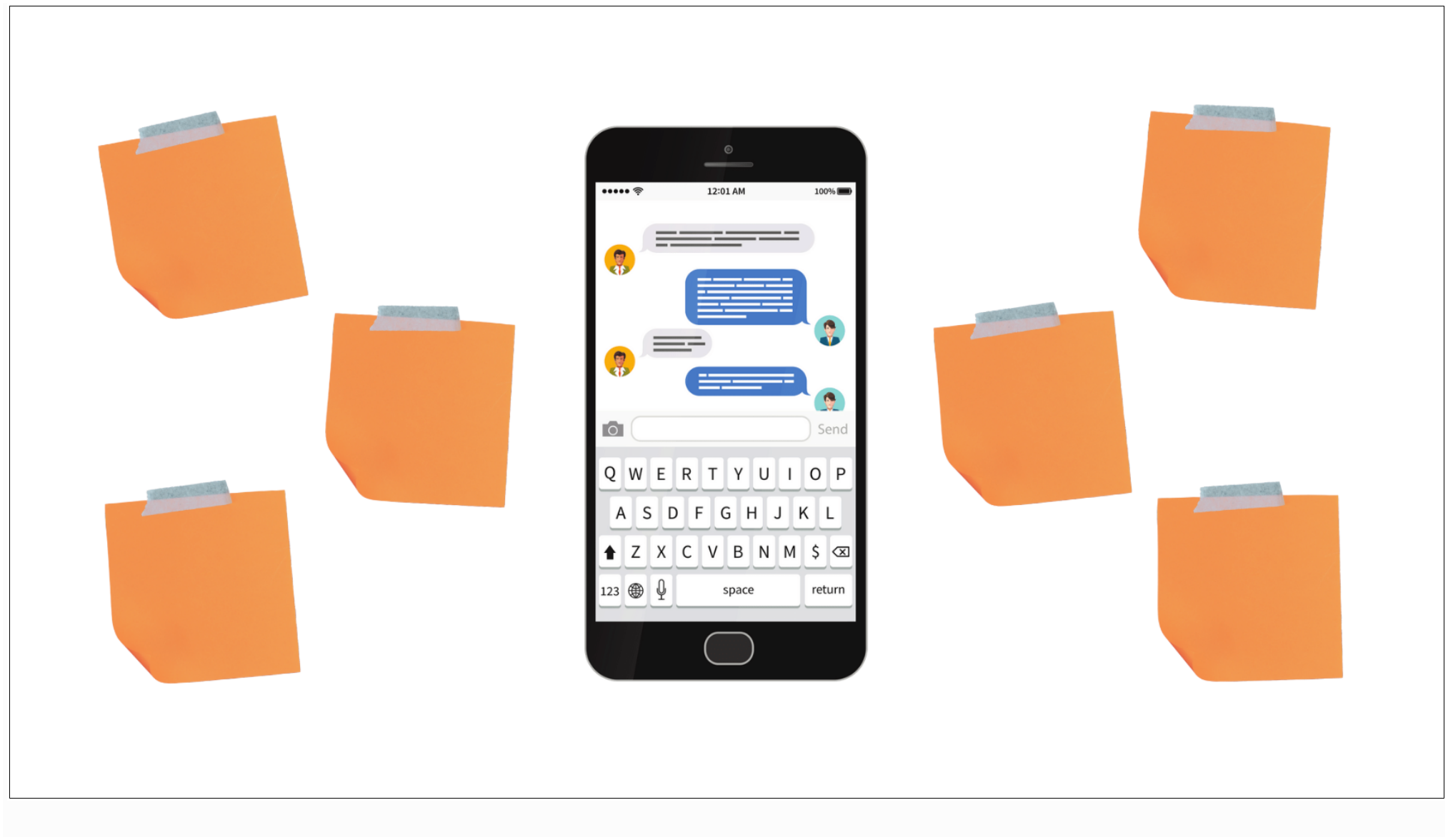

**Appendix 4:** Steps in analysis of data from the workshops with caregivers and professionals.

| **Steps** | **Aim** | **Actions** |
|---|---|---|
| 1. Meaning coding | To reflect meanings expressed | Coding of recordings from one group from both the workshop with caregivers and the workshop with professionals was conducted individually by the first author (LWHJ) and a research assistant with experiences in conducting thematic analysis. This ensured intersubjectivity of the coding process and helped form the initial coding tree. First, the whole transcript of the individual workshop was read and notes were made for initial ideas for codes for each data set. Second, coders met to compare and discuss initial codes until agreement.<br>Following this, LWHJ proceeded with the coding. Any doubts were resolved in discussion with the research assistant, SKJ or KDL. The initial coding resulted in 60 codes for the workshop with caregivers and 63 codes for the workshop with professionals. |
| 2. Meaning condensation | To reduce the volume of the text while retaining the essential meaning | All codes were then reread and condensed by the two coders through several iterations until agreement. After several rounds of coding, this reduced codes to 12 codes for the workshop with caregivers and 13 codes for the workshop with professionals.<br>These codes were then discussed to achieve further condensation and to define and name themes and subthemes that would capture the essence of data from the workshop with caregivers and professionals, respectively. |
| 3. Meaning interpretation | Developing a deeper understanding of the coded data and condensed themes | Four overall themes were defined for the workshop with caregivers and five themes for the workshop with professionals. They were described narratively and relevant quotes were chosen for presentation in the manuscript. Finally, this paper was written. |

**Appendix 5:** Matrix analysis The matrix analysis showed similarities, differences and tensions between the needs, challenges and visions for a shared digital communication intervention as perceived by caregivers and professionals. The table presents an abbreviated version of the matrix.

| **Interactional issue** | **Caregiver perspective** | **Professional perspective** | **Matrix analysis (overlap/tension)** | **Design principle** |
|---|---|---|---|---|
| **Fragmented knowledge about the child** | Repeatedly transfer information between services (want professionals to known the whole child/situation) | Lack access to comprehensive information (desire holistic understanding of the child) | Shared information gap: responsibility shifted to caregivers | *Shared child overview* |
| **Caregivers as informal coordinators** | Coordinate communication and services | Coordination lacks formal ownership | Coordination depends on caregivers and individual professionals | *Shared communication with transparent roles and responsibilities* |
| **Unequal access to support** | Support depends on family resources | Caregivers compensate for system gaps | Shared concerns about inequality | *Ensure equitable access to support* |
| **Communication depends on relationships** | Trusted professionals reduce burden | Collaboration relies on personal networks | Effective but vulnerable coordination | *Support continuity across professionals* |
| **Accessibility versus work load** | Want direct communication (easy accessibility) but fear overload for professionals involved | Want more information but fear increased workload and information overload | Better communication must not create more work | *Structured communication divided in specific tasks* |
| **Transparency and participation** | Want active involvement in all communication | Need clear boundaries and consent | Shared communication requires defined roles | *Role-based access and consent management (caregiver control)* |
| **Transitions between services** | Information is especially lost during transitions | Transitions require increased coordination | High risk points for fragmentation | *Used as transitions support and when shared planning is needed* |
| **Coordination beyond technology** | Digital communication should reduce caregiver burden | Technology cannot replace human coordination | Digital solutions require human responsibility | *Combine digital platform with human coordinator support* |

**Appendix 6:**
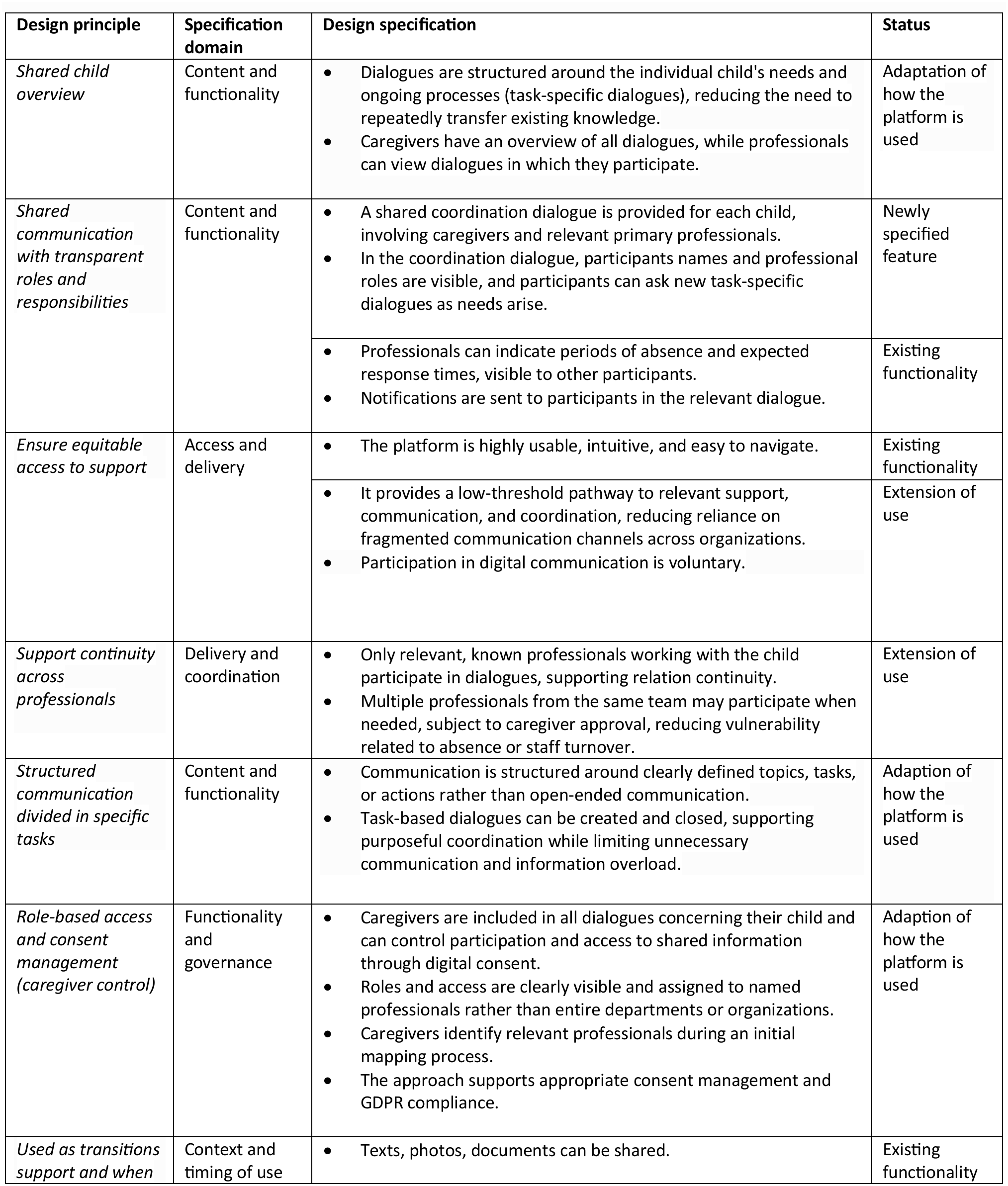
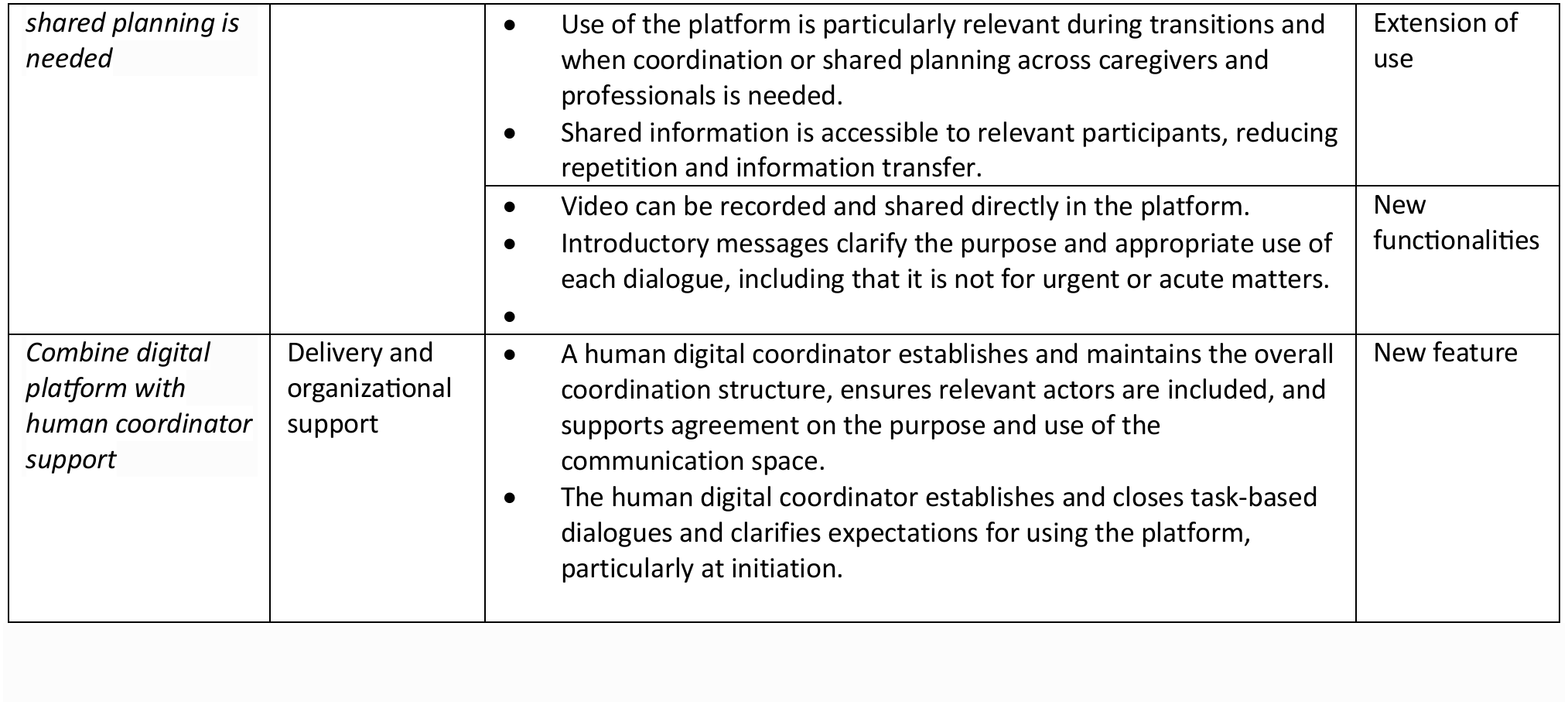
From design principles to design specifications The table shows how we moved from design principles to design specifications, the specification domain and the status of the adaptation (if it was an adaptation of intervention, an existing functionality, extension of use, or a newly specified feature).

| **Design principle** | **Specification domain** | **Design specification** | **Status** |
|---|---|---|---|
| *Shared child overview* | Content and functionality | • Dialogues are structured around the individual child's needs and ongoing processes (task-specific dialogues), reducing the need to repeatedly transfer existing knowledge.<br>• Caregivers have an overview of all dialogues, while professionals can view dialogues in which they participate. | Adaptation of how the platform is used |
| *Shared communication with transparent roles and responsibilities* | Content and functionality | • A shared coordination dialogue is provided for each child, involving caregivers and relevant primary professionals.<br>• In the coordination dialogue, participants names and professional roles are visible, and participants can ask new task-specific dialogues as needs arise. | Newly specified feature |
|  |  | • Professionals can indicate periods of absence and expected response times, visible to other participants.<br>• Notifications are sent to participants in the relevant dialogue. | Existing functionality |
| *Ensure equitable access to support* | Access and delivery | • The platform is highly usable, intuitive, and easy to navigate. | Existing functionality |
|  |  | • It provides a low-threshold pathway to relevant support, communication, and coordination, reducing reliance on fragmented communication channels across organizations.<br>• Participation in digital communication is voluntary. | Extension of use |
| *Support continuity across professionals* | Delivery and coordination | • Only relevant, known professionals working with the child participate in dialogues, supporting relation continuity.<br>• Multiple professionals from the same team may participate when needed, subject to caregiver approval, reducing vulnerability related to absence or staff turnover. | Extension of use |
| *Structured communication divided in specific tasks* | Content and functionality | • Communication is structured around clearly defined topics, tasks, or actions rather than open-ended communication.<br>• Task-based dialogues can be created and closed, supporting purposeful coordination while limiting unnecessary communication and information overload. | Adaption of how the platform is used |
| *Role-based access and consent management (caregiver control)* | Functionality and governance | • Caregivers are included in all dialogues concerning their child and can control participation and access to shared information through digital consent.<br>• Roles and access are clearly visible and assigned to named professionals rather than entire departments or organizations.<br>• Caregivers identify relevant professionals during an initial mapping process.<br>• The approach supports appropriate consent management and GDPR compliance. | Adaption of how the platform is used |
| *Used as transitions support and when shared planning is needed* | Context and timing of use | • Texts, photos, documents can be shared. | Existing functionality |
|  |  | • Use of the platform is particularly relevant during transitions and when coordination or shared planning across caregivers and professionals is needed.<br>• Shared information is accessible to relevant participants, reducing repetition and information transfer. | Extension of use |
|  |  | • Video can be recorded and shared directly in the platform.<br>• Introductory messages clarify the purpose and appropriate use of each dialogue, including that it is not for urgent or acute matters.<br>• | New functionalities |
| *Combine digital platform with human coordinator support* | Delivery and organizational support | • A human digital coordinator establishes and maintains the overall coordination structure, ensures relevant actors are included, and supports agreement on the purpose and use of the communication space.<br>• The human digital coordinator establishes and closes task-based dialogues and clarifies expectations for using the platform, particularly at initiation. | New feature |

